# Real-World Performance of the Alinity m STI Assay for the simultaneous detection of Chlamydia trachomatis, Neisseria gonorrhea, Mycoplasma genitalium, and Trichomonas vaginalis in Clinical Specimens at a Large Tertiary Hospital System in the United States

**DOI:** 10.1101/2023.12.12.23299874

**Authors:** Karen C. Rexroth, Joshua Kostera

**Author notes:** **Corresponding author:** Joshua Kostera. **Sources of support:** Abbott Molecular provided the Alinity m STI reagents for the study. **Summary** Evaluation of the Alinity m STI assay in a large US hospital system identified a higher prevalence of trichomonas and mycoplasma infections than chlamydia and gonorrhoeae infections in urogenital specimens.

## Abstract

**Background:** In the US, sexually transmitted infections (STI) are reported for Chlamydia trachomatis (CT) and Neisseria gonorrhoeae (NG) but not Trichomonas vaginalis (TV) and Mycoplasma genitalium (MG). TV and MG surveillance data in the US remains inadequate. We evaluated the performance of the fully automated, multi-plex PCR-based Alinity m STI assay, to simultaneously detect CT, NG, TV, and MG, at a large Veterans Affairs hospital system.

**Methods:** 260 urine, urogenital, and extragenital clinical specimens were tested with the Alinity m STI assay and the results were compared to those from the Abbott RealTi*m*e CT/NG assay in our lab and TV and MG assays run at an external reference laboratory.

**Results:** Alinity m STI assay results for urine specimens had an overall percent agreement (OPA) with the RealTi*m*e comparator assay of 187/187 (100.0%) for CT and 186/187 (99.5%) for NG. For 20 rectal specimens, the OPA for CT was 95% and for NG was 100%; for 13 oral specimens, OPA was 100% for CT and 92.3% for NG. Of the urine specimens sent for TV and MG testing, 100% agreement was observed between the Alinity m STI assay and TV (n=104) and MG (n=30) comparator assay results.

**Conclusions:** The Alinity m STI assay is an easy-to-use, sensitive and specific assay that allows high-throughput testing for common and undertested STI pathogens to facilitate surveillance efforts.

**KEY MESSAGES:** *What is already known on this topic:*

- Mycoplasma genitalium and Trichomonas vaginalis are sexually transmitted infections, however, global prevalence data vary between geographical regions, as these sexually transmitted infections are not included in routine screening.

*What this study adds:*

- This study aimed at assessing Trichomonas and Mycoplasma prevalence and association with a positive Chlamydia and/or Gonorrhoeae result.

*How this study might affect research, practice, or policy:*

- Including Mycoplasma genitalium and Trichomonas vaginalis testing in a routine screening STI diagnostic algorithm, with cost-effective and accurate multiplex assays can improve diagnosis to initiation of appropriate treatment.

## Introduction

In the United States, approximately 2.5 million STIs were diagnosed in 2021,^1,2^ despite recent reports declaring that Americans are having less sex in the pandemic era.^3,4^ A recent report from the World Health Organization (WHO) estimated that there were 374 million new STIs globally in 2020, with more than 1 million people infected each day with 1 of the 4 most common pathogens: Chlamydia trachomatis (CT; 129 million), Neisseria gonorrhoeae (NG; 82 million), syphilis (7.1 million), and Trichomonas vaginalis (TV; 156 million).^5^ The well-characterized bacterial pathogens CT and NG, as well as TV and Mycoplasma genitalium (MG) are recognized STI pathogens;^5-7^ however, TV and MG are not considered reportable by CDC.^8^ Undiagnosed and untreated STIs have significant and lifelong reproductive and non-reproductive health consequences, and can increase the risk of acquiring or transmitting HIV.^8-10^ Accurate diagnosis of STIs is necessary to interrupt transmission pathways and initiate timely and appropriate treatment to improve long-term health outcomes and avoid multidrug resistance.^8,11-13^

Diagnosis of STIs can be challenging, as the pathogens cause overlapping symptoms or subclinical symptoms that may be misdiagnosed or overlooked. Current guidelines recommend the use of nucleic acid amplification testing (NAAT) to screen for the reportable pathogens, CT and NG, and help reduce transmission and guide precision treatment.^14,15^ NAAT-based surveillance programs for TV and MG would allow additional detection of subclinical or asymptomatic infections and further hone treatment algorithms.^8,16,17^

To reduce the time to diagnosis and detect multiple pathogens in a single specimen, the Alinity m STI assay is a qualitative multiplex PCR-based test for the simultaneous detection of CT ribosomal RNA sequences, NG DNA sequences, TV ribosomal RNA sequences, and MG ribosomal RNA sequences extracted from endocervical swab, vaginal swab, male and female urine, oropharyngeal swab, and rectal swab specimens, and gynecological specimens preserved in ThinPrep PreservCyt^®^ Solution.^18^ The assay also amplifies and detects an endogenous human DNA sequence (cellular control, CC) and an internal control (IC) in every specimen to confirm specimen adequacy and process validity, respectively. The reporting structure of the assay releases results for any combination of CT, NG, TV, and MG along with the IC and CC results for each specimen.

In our study, we evaluated the real-world clinical performance of the Alinity m STI assay compared to other commercially available molecular assay platforms for detection of CT, NG, TV, and MG, as well as its utility for surveillance of TV and MG infection, at the Washington DC Veterans Affairs Medical Center (DC VAMC), a large tertiary hospital in the US. DC VAMC serves veterans in the District of Columbia and surrounding suburbs, Maryland, and West Virginia, which has an estimated veteran population of 765,000. The DC VAMC is an acute care hospital that includes 7 community-based outpatient clinics, a women’s clinic, infectious diseases clinic, and an emergency department; specimens included in this study were acquired at each of these locations. As Washington DC has the fifth highest STI rate in the US,^2^ the DC VAMC is a favorable site for not only evaluating the performance of the Alinity m STI assay but also examining the cross-sectional prevalence of detected STIs compared to the latest statistics released by the CDC.

## Methods

The study included 3 objectives. The first was to perform an observed sensitivity assessment using standard biological reference materials from a third-party manufacturer (Exact Diagnostics, Fort Worth, TX) to evaluate the sensitivity of the assay. The second objective was to conduct a clinical comparative evaluation in which specimens were tested with the Alinity m STI assay and other commercially available STI assays to determine result agreement. The third objective was an ad hoc stability study evaluating individual neat urine specimens confirmed positive with a STI pathogen to determine specimen integrity for up to 7 days in cold storage, either refrigerated or frozen, prior to testing with the Alinity m STI assay. All statistical analyses were completed using SAS version 9.3 or higher, or Microsoft Excel version 2108.

### Observed sensitivity assessment

To evaluate the sensitivity of the Alinity m STI assay for each of the 4 pathogens (CT, NG, TV, MG), biological reference material (Exact Diagnostics) for each organism was used. Materials were stored and handled according to the labeling instructions provided by the manufacturer. The material contains whole intact biological organism in a synthetic matrix and contains preservative that is value assigned using digital droplet PCR with assigned units in copies/mL. Alinity m multi-Collect Specimen Collection Kit stabilization buffer was used to make up a bulk diluent, which was then used to prepare multiple panels where the reference material was spiked to create diluted titers for each of the 4 pathogens. A range-finding study was performed for each of the pathogens to determine the observed sensitivity of the assay. This was followed by testing 10 replicates at the 2 lowest range-finding titer levels at which the assay could detect 95% or greater of replicates.

### Clinical comparative evaluation

A total of 260 deidentified adult urogenital and extragenital remnant clinical specimens were included in this study (Figure 1), which was determined to be exempt by the DC VAMC IRB. Specimens were acquired after obtaining informed consent from consecutive inpatients and outpatients at the DC VAMC. Specimens were collected in accordance with institutional standard practices and procedures as part of routine patient care and consisted of male and female urine and rectal, oropharyngeal, vaginal, and endocervical swabs. Additional gynecological specimens acquired for the study were collected in ThinPrep PreservCyt^®^ and stored according to the manufacturer instructions.

**Figure 1.**
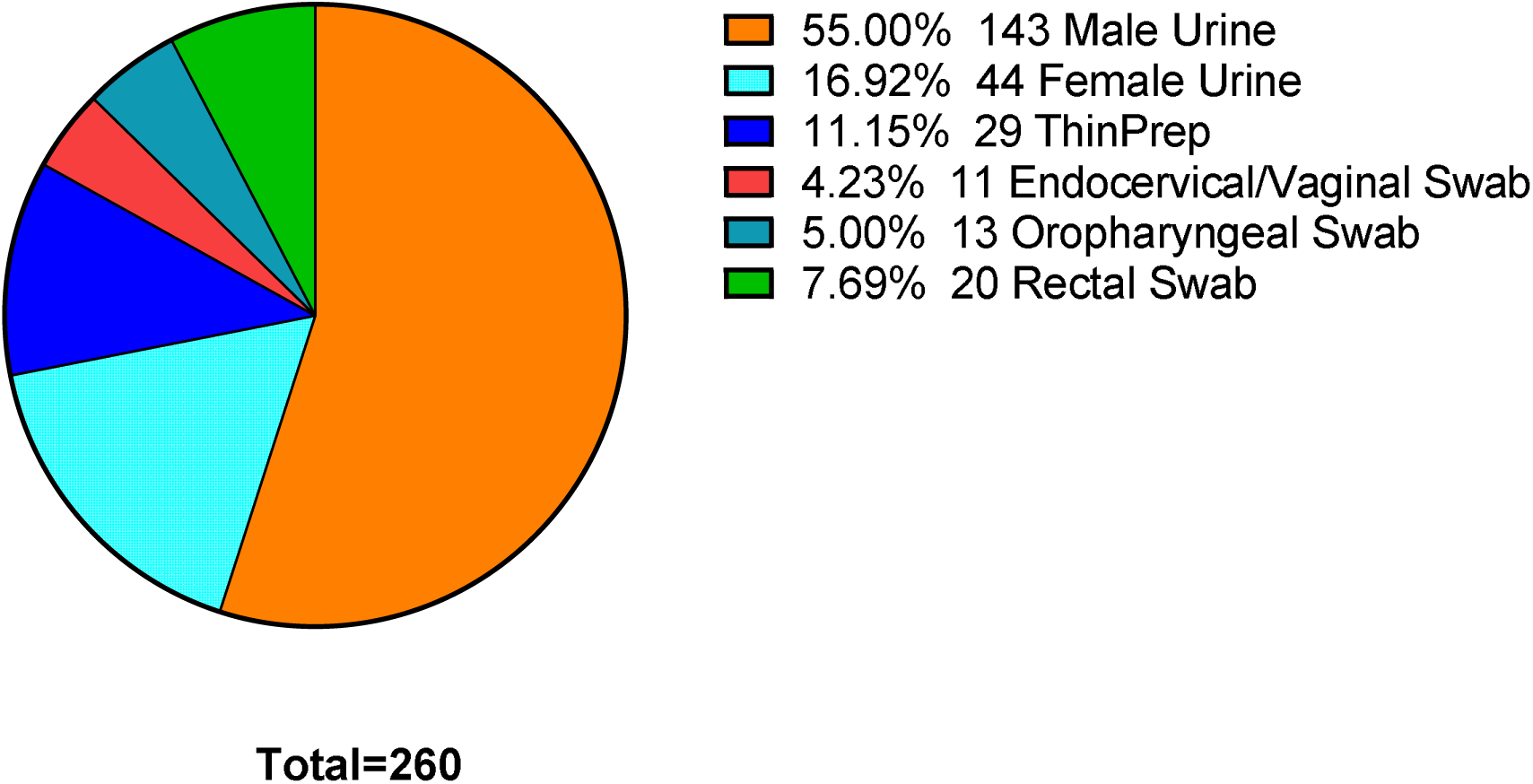
Stratification of specimen types included in the clinical comparative study.

Clinical swab specimens included in the study were dual collected using the Abbott multi-Collect Specimen Collection Kit (Abbott Kit) and the Alinity m multi-Collect Specimen Collection Kit (Alinity Kit). The 2 collection kits are intended for the collection and transportation of male and female urine specimens, endocervical swab specimens, vaginal swab specimens, oropharyngeal swab specimens, and rectal swab specimens to stabilize nucleic acid for testing with the Alinity m STI assay. Urine specimens sent to the laboratory were split between the 2 collection kits, and approximately 2 mL of specimen was aseptically transferred to the collection kit tube according to the manufacturer’s instructions. All specimens once added to the respective collection devices were stored refrigerated at 2-8°C for up to 14 days or frozen at -20°C for up to 60 days. The Alinity kit specimens were tested with the Alinity m STI assay and Abbott kit specimens were tested with the Abbott RealTi*m*e CT/NG assay (Abbott Laboratories). The clinical evaluation was conducted using 3 lots of amplification reagents, controls, and bulk specimen extraction reagents. Alinity m STI results for CT and NG were compared to the Abbott RealTi*m*e CT/NG assay results. Alinity m STI results for TV and MG were compared to those from specimens sent to an external reference laboratory service (Labcorp, Burlington, NC). Specimens that were sent to the external reference laboratory service were also collected in the Aptima Transport tube. The external laboratory service tests TV using TMA technology and MG using the Sureswab^®^ Mycoplasma/Ureaplasma Panel. Extragenital specimen testing was previously self-validated for use with the Abbott RealTi*m*e CT/NG assay.

### Urine specimen stability assessment

To evaluate neat urine specimen stability prior to processing, patients followed the clean catch urine collection procedure using a sterile collection container. Specimens that were individually identified as positive for CT, NG, TV, and MG were split into 2 aliquots. One aliquot was refrigerated at 4°C and the other was aliquoted into 4 transport tubes containing no preservative buffer and capped. These were stored at frozen at -20°C. Individual specimen aliquot tubes were used to mitigate the freeze-thaw impact on stability of the specimens; the impact of freeze-thaw cycles on specimen stability was not assessed in this investigation. The stability testing schedule included test points on day 1, 3, 5, and 7 to simulate a delayed specimen testing scenario. The cycle number (CN) for each specimen was recorded for each test point to determine the impact of storage temperature and time on Alinity m STI assay performance.

## Results

### Alinity m STI is sensitive for detection of all 4 pathogens

Prior to testing clinical specimens, an observed sensitivity assessment was performed to determine the overall sensitivity of the Alinity m STI assay. The sensitivity assessment was conducted using 1 lot of amplification reagents, controls, and bulk specimen extraction reagents. An initial range finding experiment was completed (Supplemental Table s1), followed by a final sensitivity confirmation. The limit of detection of the Alinity m STI assay was 5 copies/mL for CT, 50 copies/mL for NG, 50 copies/mL for TV, and 900 copies/mL for MG. Table 1 shows the final observed sensitivity results as well as calculated conversion estimates to units presented in the Alinity m STI assay package insert.^18^

**Table 1.**
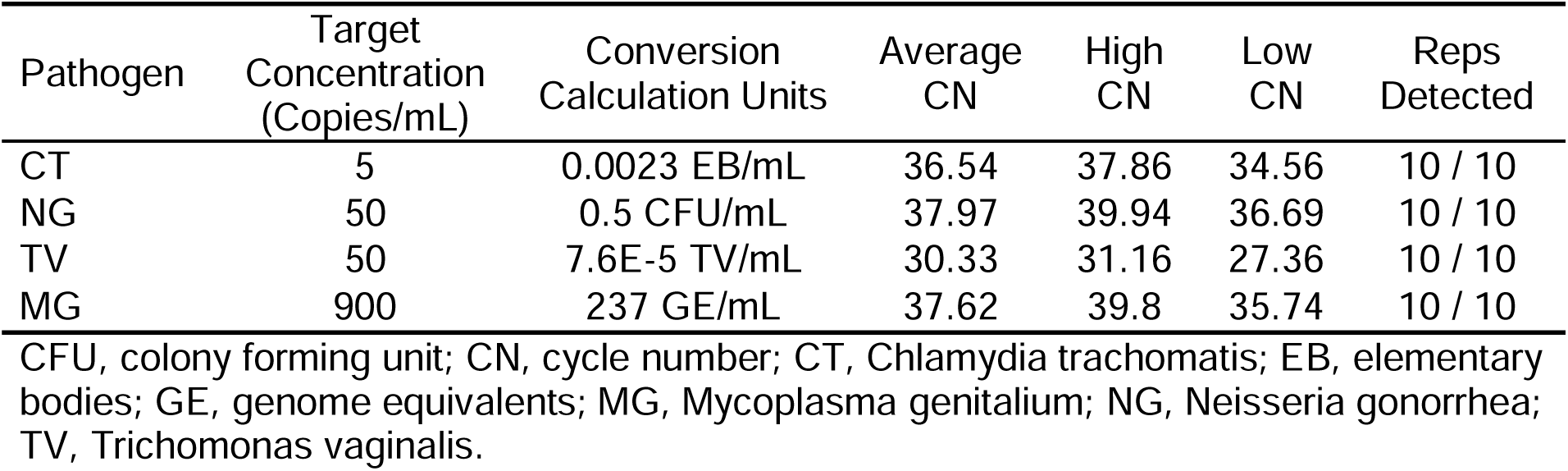
Final Observed Sensitivity of the Alinity m STI Assay.

| Pathogen | Target<br>Concentration<br>(Copies/mL) | Conversion<br>Calculation Units | Average<br>CN | High<br>CN | Low<br>CN | Reps<br>Detected |
| --- | --- | --- | --- | --- | --- | --- |
| CT | 5 | 0.0023 EB/mL | 36.54 | 37.86 | 34.56 | 10 / 10 |
| NG | 50 | 0.5 CFU/mL | 37.97 | 39.94 | 36.69 | 10 / 10 |
| TV | 50 | 7.6E-5 TV/mL | 30.33 | 31.16 | 27.36 | 10 / 10 |
| MG | 900 | 237 GE/mL | 37.62 | 39.8 | 35.74 | 10 / 10 |

CFU, colony forming unit; CN, cycle number; CT, Chlamydia trachomatis; EB, elementary bodies; GE, genome equivalents; MG, Mycoplasma genitalium; NG, Neisseria gonorrhea; TV, Trichomonas vaginalis.

### Alinity m STI assays produce comparable clinical results across specimen types

For clinical specimens, Alinity m STI results for CT and NG were compared to Abbott RealTi*m*e CT/NG assay results; Alinity m STI results for TV and MG results were compared to those received from an external reference laboratory service for NAAT testing. Reflecting the current state of STI testing in the US and the real-world design application of the study, not all specimens included in our evaluation were ordered for TV and MG testing. Based on the patient risk profile and clinical presentation, specimens were ordered and sent out for TV and MG testing at the direction of the patient provider. For the subset of specimens for which comparator testing results were available, Alinity m STI results were compared to external assay testing for 104 specimens for TV and 30 specimens for MG.

Using the Abbott RealTi*m*e CT/NG assay and external laboratory service tests for TV and MG as a reference, the Alinity m STI assay produced an OPA ≥92.3% for extragenital specimens and ≥ 99.5% for urine specimens. The clinical OPA, NPA, and PPA for the Alinity m STI assay across sample types are presented in Table 2.

**Table 2.** Clinical Sample Agreement.

| Rectal CT |  | M2000 |  | OPA | PPA | NPA |
| --- | --- | --- | --- | --- | --- | --- |
|  |  | POS | NEG |  |  |  |
| Alinity m | POS | 6 | 0 | 95.0% | 85.7% | 100% |
|  | NEG | 1 | 13 | (76.39,99.11) | (48.69,97.43) | (77.19,100.00) |
| Rectal NG |  | M2000 |  | OPA | PPA | NPA |
|  |  | POS | NEG |  |  |  |
| Alinity m | POS | 5 | 0 | 100% | 100.0% | 100.0% |
|  | NEG | 0 | 13 | (82.41,100.00) | (56.55,100.00) | (77.19,100.00) |
| Oral CT |  | M2000 |  | OPA | PPA | NPA |
|  |  | POS | NEG |  |  |  |
| Alinity m | POS | 0 | 0 | 100.0% | . | 100.0% |
|  | NEG | 0 | 13 | (77.19,100.00) | (66.69,98.63) | (77.19,100.00) |
| Oral NG |  | M2000 |  | OPA | PPA | NPA |
|  |  | POS | NEG |  |  |  |
| Alinity m | POS | 4 | 0 | 92.3% | 80% | 100.0% |
|  | NEG | 1 | 8 | (66.69,98.63) | (37.55,96.38) | (67.56,100.00) |
| Urine CT |  | M2000 |  | OPA | PPA | NPA |
|  |  | POS | NEG |  |  |  |
| Alinity m | POS | 13 | 0 | 100.0% | 100.0% | 100.0% |
|  | NEG | 0 | 174 | (97.99,100.00) | (77.19,100.00) | (97.84,100.00) |
| Urine NG |  | M2000 |  | OPA | PPA | NPA |
|  |  | POS | NEG |  |  |  |
| Alinity m | POS | 10 | 1 | 99.5% | 100.0% | 99.4% |
|  | NEG | 0 | 176 | (97.03,99.91) | (72.25,100.0) | (96.87,99.90) |
| Urine TV |  | Aptima |  | OPA | PPA | NPA |
|  |  | POS | NEG |  |  |  |
| Alinity m | POS | 9 | 0 | 100.0% | 100.0% | 100.0% |
|  | NEG | 0 | 177 | (97.98,100.00) | (70.09,100.00) | (97.88,100.00) |
| Urine MG |  | SureSwab |  | OPA | PPA | NPA |
|  |  | POS | NEG |  |  |  |
| Alinity m | POS | 21 | 0 | 100.0% | 100.0% | 100.0% |
|  | NEG | 0 | 166 | (97.99,100.00) | (84.54,100.00) | (97.74,100.00) |

CT, Chlamydia trachomatis; MG, Mycoplasma genitalium; NG, Neisseria gonorrhea; NPA, negative percent agreement; OPA, overall percent agreement; PPA, positive percent agreement; TV, Trichomonas vaginalis.

Although the sample size was smaller (n=33) for extragenital specimens than for urogenital specimens (n=227), the observed prevalence for CT (18.2%) and NG (27.3%) in extragenital specimens was much higher than the mono-infection urogenital prevalence for CT (5.0%) and NG (5.0%). Co-infections were also detected in the study cohort; 9.0% of rectal specimens and 1.0% of urine specimens were positive for both CT and NG with the Alinity m STI assay and correlated 100% with the comparator assay. One (0.20%) urine specimen was positive for CT and MG with the Alinity m STI assay, with the CT result confirmed on the Abbott RealTi*m*e CT/NG assay (this specimen was not ordered for MG testing). One (3.0%) rectal specimen was positive for NG and MG with the Alinity m STI assay, with the NG result confirmed on the Abbott RealTi*m*e CT/NG assay (this specimen was not ordered for MG testing).

In the clinical specimen evaluation data set, 3 discrepant results were observed between the Alinity m STI assay and reference tests (Abbott RealTi*m*e CT/NG or external laboratory services TV and MG tests). The 3 descrepant results were 1 urine and 1 oral specimen discrepant for NG and 1 rectal specimen descrepant for CT results. All 3 specimens had very high CNs, indicating very low levels of target pathogen. All specimens generated valid results on the Alinity m STI assay (valid IC and CC results) with no inhibition observed, including for extragenital rectal and oropharyngeal swab specimen types (data not shown).

The number and sources of clinical specimens included in this study are summarized in Table 3 along with the Alinity m STI assay testing results. Twenty-nine specimens in ThinPrep PreservCyt^®^ were tested with the Alinity m STI assay; however, all specimens were negative for CT, NG, TV, and MG. Of note, the combined total prevalence of non-reportable pathogens TV and MG (n=31) was observed to be 15.2% in the urogenital study population compared to the combined prevalence of CT and NG (n=25), which was observed to be 10.1%.

**Table 3.** Alinity m STI Assay Results by Clinical Specimen Type.

| Clinical Specimen Source | Total Tested | CT+ | NG+ | TV+ | MG+ |
| --- | --- | --- | --- | --- | --- |
| Male Urine | 143 | 12 | 11 | 7 | 18 |
| Female Urine | 44 | 1 | 0 | 2 | 3 |
| Rectal | 20 | 7 | 6 | 0 | 1 |
| Oropharyngeal | 13 | 0 | 4 | 0 | 0 |
| Endocervical / Vaginal | 11 | 0 | 1 | 0 | 1 |
| ThinPrep PreservCyt | 29 | 0 | 0 | 0 | 0 |
| Total | 260 | 25 | 27 | 11 | 23 |

CT, Chlamydia trachomatis; MG, Mycoplasma genitalium; NG, Neisseria gonorrhea; TV, Trichomonas vaginalis.

### Urine specimens are stable for up to 7 days at 4°C or *-20°C*

The objective of this study component was to investigate potential influence of storage temperature and time on the stability of neat urine specimens, based on the CN values from the Alinity m STI assay.

All CN data points for both refrigerated and frozen storage conditions were within 2 SD of the average CN for all specimens in the study, indicating that there were no statistically significant data points that could suggest significant degradation of the specimens, data shown in supplemental Table s2.

## Discussion

Our clinical findings are consistent with those of another recent study^20^ comparing the clinical performance of the Alinity m STI assay with Abbott RealTi*m*e CT/NG assay, which showed similar agreement for detecting CT, NG, and MG in patient specimens. A separate multicenter study also showed high correlations between the Alinity m STI assay and various comparator assays.^21^ In our study, across all specimen types in the entire study cohort, 25.0% (n=65/260) of patient specimens were found to have either a mono or co-infection STI. STIs that are not accurately diagnosed will continue to challenge public health causing significant and serious long-term health consequences for patients.

Historically, MG has not been routinely tested at DC VAMC; as all 4 pathogens can be simultaneously detected by the Alinity m STI assay, it is notable that MG was observed to have the highest positivity rate compared to the other STI pathogens in our specimen cohort. 12.6% of male urine specimens were positive for MG, compared to 8.4% for CT and 7.7% for NG. For female urine specimens in our study, MG was detected in 6.8% of specimens, also higher than that detected for CT (2.27%), and no cases of NG were detected. Although 187 urogenital specimens were not ordered for MG testing, 16 (∼10%) of those were positive for MG and negative for CT, NG, and TV on the Alinity m STI assay. All CT and NG results were confirmed with comparator testing and 109 out of the 260 specimens were ordered and confirmed for TV with comparator testing. MG infections are generally asymptomatic in both men and women, contributing up to 35% of non-chlamydial non-gonococcal urethritis in men and linked to cervicitis and pelvic inflammatory disease in women.^6^ Clinical treatment failures and antimicrobial-resistant MG are a significant concern, and rising incidence rates of unknown magnitude have complicated the clinical management of this infection. The consequences of an undiagnosed and untreated MG infection can be severe; our findings support the need for STI multi-plex testing to include MG.

Globally, TV is the most common, curable non-viral STI that is often associated with serious health consequences including preterm birth, low-birthweight infants, and infertility, as well as increasing the risk for acquisition and transmission of human immunodeficiency virus (HIV).^5,8^ The epidemiology of TV in men is less delineated than for women, for the male urine specimens included in our study, we observed a 4.9% positivity rate for TV. Risk factors for TV in women include older age, race, impoverished socioeconomic status, and multiple sexual partners.^10^ In our study, 4.5% of female urine specimens were positive for TV. At present, neither TV nor MG meet all the criteria for confirmed cases to be reported to the CDC, including for women who are pregnant.

Our study findings support the use of NAATs for testing and surveillance of STI pathogens such as MG and TV. Simultaneous detection of the 4 major STI pathogens by the Alinity m STI assay would simplify the implementation of MG/TV surveillance programs in health systems that already report NG and CT infections. TV and MG surveillance programs can lead to evidence-driven changes in the STI diagnostic algorithm, with cost-effective and accurate multiplex assays moving patients quickly from diagnosis to initiation of appropriate treatment.

Infections at extragenital sites are typically asymptomatic and latent, making clinical diagnosis challenging in the absence of a sensitive and specific molecular assay. Extragenital specimens were included in our study to evaluate the performance of the Alinity m STI assay with oropharyngeal and rectal swabs. Extragenital testing is not always part of routine STI screening algorithms, although recent studies have highlighted the STI incidence rate at the oropharynx and rectum, leading to a shift in practice to include screening of extragenital sites.^19^ In total, 33 extragenital specimens were submitted for testing in our study. Four out of 13 oropharyngeal specimens (30.8%) were positive for NG. Three out of 20 rectal specimens (15.0%) were positive for both CT and NG, 5 (25.0%) were positive for NG, 4 (20.0%) were positive for CT, and 1 (5.0%) was positive for both NG and MG. The extragenital specimens included in this study were primarily collected from male patients (n=32), with only 1 extragenital specimen from a female patient. Overall, men who have sex with men (MSM) demonstrate a higher prevalence of extragenital infection compared to women and men who have sex with women;^22^ however, extragenital STI testing should be considered for both men and women.^22^ An increased risk of HIV acquisition is a potential consequence of untreated extragenital infections, which can serve as a potential reservoir for transmission specifically in patients with repeat gonococcal and chlamydial rectal infections.^23^ Surveilling the frequency of STI will inform how trends in screening programs change over time as HIV PrEP access expands and STI screening programs become more widely adopted among PrEP users.^22^

Staffing shortages and specimen transport logistical challenges can introduce unique impediments that reduce laboratory testing efficiency. Patient specimen hold times can vary prior to processing into the appropriate collection device, tube, or media that is utilized with the testing methodology or system to analyze the specimen. Furthermore, current guidelines in the literature are contradictory for urine storage temperatures and duration prior to analysis. The findings from our assessment suggest that correctly collected neat urine specimens intended for STI testing that cannot be processed immediately maintain integrity when stored refrigerated or frozen for up to 7 days prior to processing. Differences in bio-metabolic profiles that can impact specimen integrity were not examined.

## Data Availability

All data produced in the present study are available upon reasonable request to the authors

## Acknowledgements

The authors would like to thank Yan Zhang, PhD, for assistance with the statistical analysis. Stacey Tobin, PhD, provided editorial support for manuscript preparation, with compensation from Abbott Laboratories.

## Supplemental

**Table s1.** Observed Sensitivity Range-Finding Results for the Alinity m STI Assay.

| Pathogen | Levels Tested<br>(Copies / mL) | Average CN | Reps<br>Detected |
| --- | --- | --- | --- |
| CT | 300 | 35.54 | 3 / 3 |
|  | 150 | 35.5 | 3 / 3 |
|  | 75 | 35.96 | 3 / 3 |
|  | 20 | 37.45 | 3 / 3 |
|  | 10 | 37.7 | 3 / 3 |
|  | 5 | 38.59 | 2 / 3 |
| NG | 500 | 32.51 | 3 / 3 |
|  | 250 | 33.98 | 3 / 3 |
|  | 100 | 35.45 | 3 / 3 |
|  | 50 | 37.00 | 3 / 3 |
|  | 40 | 37.93 | 2 / 3 |
| TV | 150 | 32.00 | 3 / 3 |
|  | 75 | 31.85 | 3 / 3 |
|  | 50 | 31.42 | 3 / 3 |
|  | 25 | N / D | 0 / 3 |
|  | 10 | N / D | 0 / 3 |
| MG | 1700 | 28.53 | 3 / 3 |
|  | 1500 | 29.27 | 3 / 3 |
|  | 1000 | 30.37 | 3 / 3 |
|  | 900 | 29.57 | 0 / 3 |
|  | 750 | 38.95 | 2 / 3 |

CN, cycle number; CT, Chlamydia trachomatis; MG, Mycoplasma genitalium; NG, Neisseria gonorrhea; TV, Trichomonas vaginalis.

**Table s2.** Neat Urine Stability Summary.

| Pathogen | CT |  | NG |  | TV |  | MG |  |
| --- | --- | --- | --- | --- | --- | --- | --- | --- |
| Baseline CN | 27.78 |  | 24.5 |  | 21.75 |  | 27.24 |  |
|  | -20°C | 4°C | -20°C | 4°C | -20°C | 4°C | -20°C | 4°C |
| Day 1 CN | 27.49 | 27.62 | 24 | 26.49 | 23.61 | 22.42 | 27.75 | 27.23 |
| Day 3 CN | 27.96 | 27.75 | 24.25 | 27.13 | 25.1 | 23.37 | 28.69 | 27.55 |
| Day 5 CN | 27.36 | 27.62 | 25.24 | 27.63 | 24.42 | 24.89 | 27.86 | 26.99 |
| Day 7 CN | 27.62 | 27.29 | 25.57 | 27.43 | 26.73 | 25.82 | 28.86 | 27.5 |
| Average CN | 27.61 | 27.57 | 24.77 | 27.17 | 24.97 | 24.13 | 28.29 | 27.32 |
| SD | 0.26 | 0.20 | 0.76 | 0.50 | 1.32 | 1.52 | 0.57 | 0.26 |
| 2 SD | 0.52 | 0.39 | 1.52 | 1.00 | 2.65 | 3.04 | 1.13 | 0.52 |
| CFU, colony forming unit; CN, cycle number; CT, Chlamydia trachomatis; MG, Mycoplasma genitalium; NG, Neisseria gonorrhea; SD, standard deviation; TV, Trichomonas vaginalis. |  |  |  |  |  |  |  |  |

